# Real-time Hearing Threshold Determination of Auditory Brainstem Responses by Cross-correlation Analysis

**DOI:** 10.1101/19003301

**Authors:** Haoyu Wang, Bei Li, Yan Lu, Kun Han, Haibin Sheng, Jialei Zhou, Yumeng Qi, Xueling Wang, Zhiwu Huang, Lei Song, Yunfeng Hua

## Abstract

Auditory brainstem response (ABR) serves as an objective indication of auditory perception at given sound level and is nowadays widely used in hearing function assessment. Despite efforts for automation over decades, hearing threshold determination by machine algorithm remains unreliable and thereby still rely on visual identification by trained personnel. Here, we described a procedure for automatic threshold determination that can be used in both animal and human ABR tests. The method terminates level averaging of ABR recordings upon detection of time-locked waveform through cross-correlation analysis. The threshold level was then indicated by a dramatic increase in the sweep numbers required to produce “qualified” level averaging. A good match was obtained between the algorithm outcome and the human readouts. Moreover, the method varies the level averaging based on the cross-correlation, thereby adapting to the signal-to-noise ratio of single sweep recordings. These features empower a robust and fully automated ABR test.

## Introduction

The auditory brainstem responses (ABRs) are brain electrical potential changes due to synchronous neuronal activities evoked by suprathreshold acoustic stimuli (Jewett et al., 1970). These responses are detectable using non-invasive surface electrodes placed on the scalp of test subject and thereby widely used for hearing function assessment. In rodent and cat, typical ABR waveform is composed of initial five peaks in the early onset of sound evoked potentials, representing synchronous activities arising from projections along the auditory ascending pathway including auditory nerve, cochlear nucleus, superior olivary complex, lateral lemniscus and inferior colliculus, respectively (Henry, 1979; Melcher et al., 1996), whereas human has slightly different peak generators as demonstrated with intracranial recordings (Moller and Jannetta, 1983) and neuromagnetic responses (Parkkonen et al., 2009). Thus, ABR wave latencies and amplitudes provide clinical-significant information, for instance site of lesions or tumors in the auditory system (Lewis et al., 2015; Roeser et al., 2007) based on how the properties of ABR waveform are altered.

Although the ABR test itself is an objective measurement, the determination of threshold involves human interpretation of the ABR waveform. The readout of ABR threshold requires a trained personnel to supervise waveform recognition, which is labor-demanding. Besides, such interpretations oftentimes are subjective and may introduce errors that vary from person to person depending on his/her skill and experience, especially in cases with atypical waveform or with high background noise (Vidler and Parkert, 2004). In order to accurately detect mild hearing threshold elevation in the diagnosis of progressive hearing loss (Barreira-Nielsen et al., 2016), hidden hearing loss (Kujawa and Liberman, 2009; Mehraei et al., 2016; Ridley et al., 2018; Sergeyenko et al., 2013), age-related hearing loss (Gates and Mills, 2005; Sergeyenko et al., 2013) and tinnitus (Bramhall et al., 2018; Castaneda et al., 2019), unbiased automatic approaches with high precision and reliability are essential, particularly when screening is involved. Over decades, many attempts were made to automate the procedure, including (1) quantification of the waveform similarity by comparing either existing templates (Davey et al., 2007; Elberling, 1979; Valderrama et al., 2014) or based on features learned by artificial neural network from human annotation (Alpsan and Ozdamar, 1991; McKearney and MacKinnon, 2019); (2) quantification of the waveform stability by cross-correlation function between single-sweeps (Bershad and Rockmore, 1974; Weber and Fletcher, 1980), interleaved responses (Berninger et al., 2014; Xu et al., 1995) or responses at adjacent stimulus levels (Suthakar and Liberman, 2019); (3) the ‘signal quality’ through scoring procedures like F-ratios (Cebulla et al., 2000; Don and Elberling, 1994; Elberling and Don, 1984; Sininger, 1993). However, due to heterogeneity in inter-subject waveform and signal-to-noise-ratio (SNR) introduced by variations in test subject conditions, electrode placement/impedance, as well as acquisition settings, the accurate threshold determination is only possible under a narrow range of experimental settings, which hampers direct comparisons of ABR results across laboratories.

In this study, we proposed an automated approach for real-time ABR threshold determination. Instead of using a prefixed sweep number, level averaging was instructed based on the outcomes of cross-correlation analysis during ongoing sweep acquisition. Near-threshold stimuli feature a sharp increase in the end sweep number upon the detection of time-locked response. We further explored the potential of using this feature for the threshold determination in human subjects and the algorithm outcomes were validated by the human readouts of the same ABR level representations.

## Results

### Termination of On-going Averaging by Cross-correlation Analysis

Auditory brainstem responses (ABRs) registered by surface electrodes are embedded in high-level background activities and system noise. Smooth baseline and clear waveform, if present, are obtained after averaging over hundreds of sweeps. The number for averaging, however, depends on the amplitude of the evoked response, which varies between test subjects due to variations in, for instance, skull sizes, electrode impedances and placement that determine the distance from the peak generator and the vector projection to the electrodes. Within one recording session, these experimental parameters are usually fixed and the baseline activities at different stimulus levels are comparable, allowing signal-to-noise-ratio (SNR) quantification. It is expected that strong responses from high level stimuli quickly reach confident SNR level, while weak response evoked by low level stimulus requires more averaging and for subthreshold recordings the SNR cannot be improved by level averaging (**Figure 1A**).

**Figure 1.**
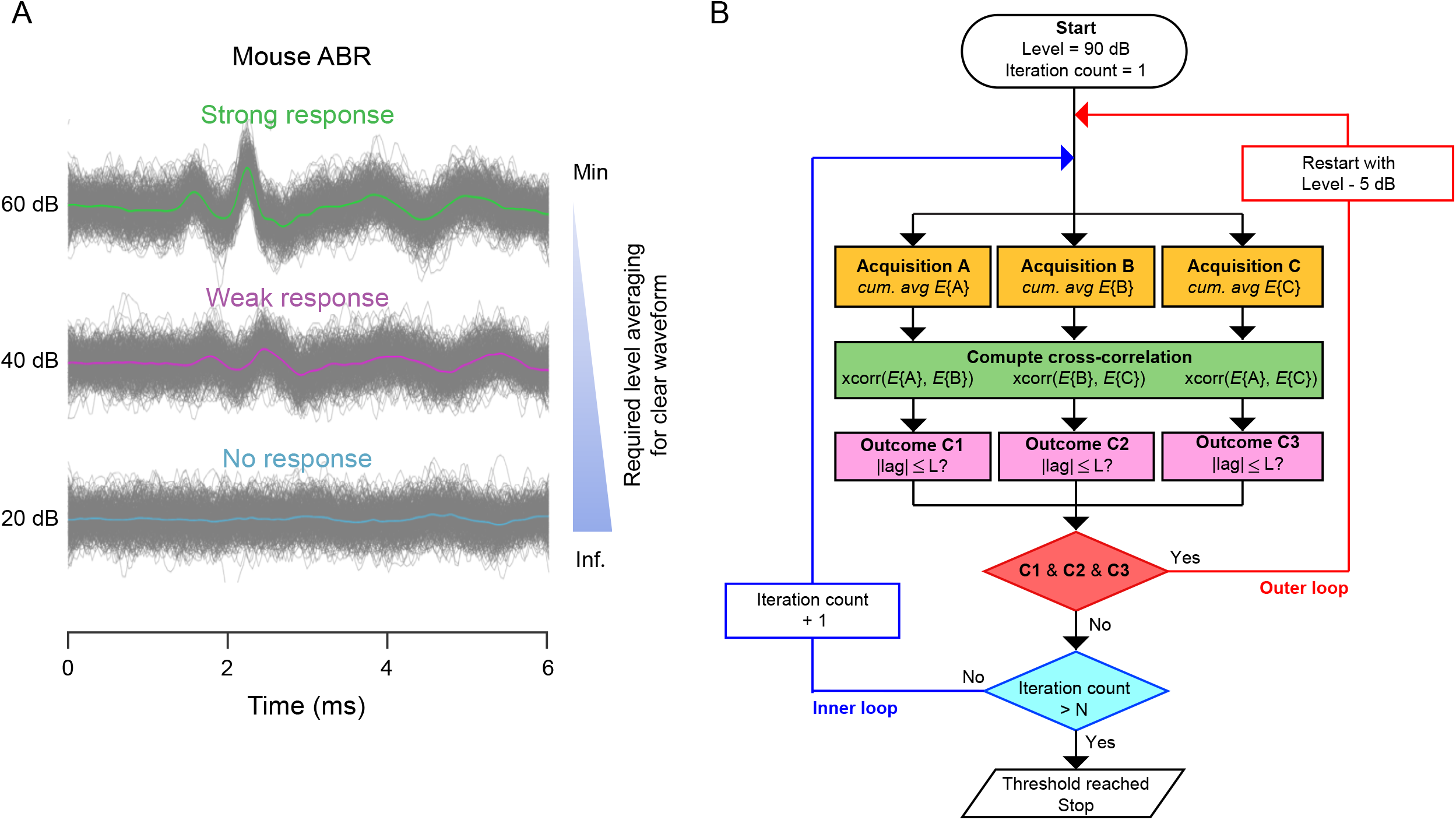
Principle and algorithm design for automated threshold determination. (A) Typical level representation of ABRs. To obtain stable waveform, one needs less level averaging of sweep recordings at strong stimulus (green) than that of weak response (magenta), whereas level averaging cannot improve the SNR of subthreshold recordings containing no response (blue). (B) Algorithm flowchart. The test starts with the highest stimulus level (e.g., 90 dB SPL for mouse ABR) and a reset iteration count. Recorded sweep batches (e.g., 60 sweeps for mouse ABR) are cumulatively averaged in three data buffers (yellow boxes, *E*{A}, *E*{B} and *E*{C}). Cross-correlation operation (*xcorr*) is carried out in each two out of three group averages (green boxes), which yields three CC peaks and their corresponding signal lags. If the absolute lag is less than the allowed value (L ≤ 2 data points for mouse ABR), the outcome (magenta boxes, C1–C3) returns true, suggesting a positive response. In cases of all three positive outcomes (red box), the procedure descends to lower stimulus level (red line, the outer loop), otherwise the same stimulus level will be repeated with more sweeps (the inner loop, blue line). An iteration upper limit (blue box, e.g., N = 7 for mouse ABR) is implemented to jump out the inner loop upon subthreshold stimulus and flag no response.

Based on above notions, we designed a procedure to test whether the change in sweep number is required for the average response at different stimulus levels to reach a certain SNR level. Such change can produce unbiased confidant threshold reading. In detail, at a given stimulus level recorded sweeps are loaded into three memory buffers (**Figure 1B**, yellow boxes) and cross-correlation coefficients (CCs) are computed between two out of three group averages (green boxes). If time-locked response, irrespective of wave latencies and shapes, is present, two group averages overlap with each other at time zero, and thereby the obtained CC peak is found within a small time shift (L) from zero (magenta boxes). Three parallel runs (red box) can effectively reject false positives caused by overlapped random noise with similar peak latencies. Next, the correlation analysis is iterated with increasing sweep numbers (the inner loop). Once the correlation analysis identifies a time-locked response, the iteration will end with sweep number noted. Upon the absence of a time-locked response, the upper limit for iteration count (N) is used to terminate nonproductive attempts. Finally, the outer loop is implemented to scan the threshold response with decreasing stimulus levels and the stop command is triggered upon two consecutive levels that reached the iteration limit.

### ABR Threshold Determination in Mouse

To test whether the proposed algorithm could determine the ABR threshold reliably, we recorded from ten mice (three wild-type adult C57BL/6 mice of normal hearing, two wild-type adult CBA mice experienced noise exposure and five telomerase knock-out mice with early-onset hearing loss; data are pooled in this study) single-sweep ABR sets starting from 90 dB (sound pressure level, SPL) to 0 dB with a step size of 5 dB. The raw data were corrected for baseline fluctuations through a smoothing spline fit before being processed by the algorithm.

Figure 2 presents a walkthrough of the procedure. First, an example of level series is plotted (**Figure 2A**) and the visually identified threshold denoted at 30 dB SPL with an asterisk. In the algorithm, three level averages (**Figure 2B**) were used to compute the correlation coefficient (CC). The changes in CC peak amplitude (**Figure 2C**) and the corresponding signal lag (**Figure 2D**) upon different stimulus levels were then obtained to further explore whether the two derivatives can predict the threshold. Upon reducing stimulus strength, a monotonic decrease was observed in the CC peak amplitudes, whereas near-threshold levels feature a sudden jump from small absolute lags (≤ 2 data points or less than ± 0.082 ms for suprathreshold levels) to big absolute lags with large variation at subthreshold levels. This result suggested that the signal lag is a better response detector at near-threshold levels, thus justified its use in the algorithm to demarcate the threshold boundary.

**Figure 2.**
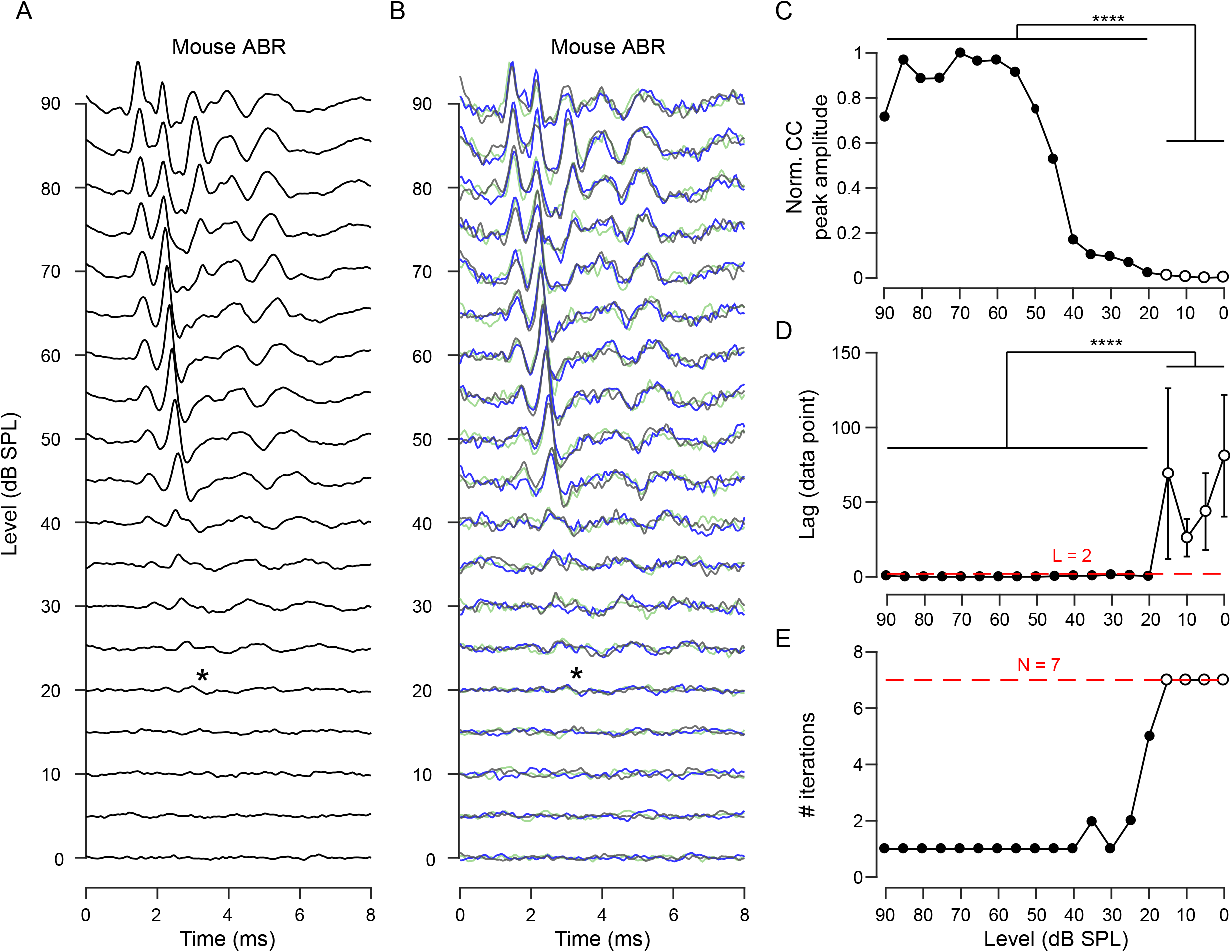
Cross-correlation analysis of mouse ABR. (A) Example level representation (averaging over 420 sweeps) of a mouse. The visually identified threshold level was 20 dB SPL (asterisk). (B) Group averages generated for cross-correlation analysis. (C) Plot of the obtained CC peak amplitude versus the level series. (D) Plot of signal lag at the CC peak versus the level series. At suprathreshold levels (dots, ≥ 20 dB), small signal lags were consistent across three parallel runs (0.33 ± 0.56 data points, mean ± s.d.), while at subthreshold levels (cycles, < 30 dB) large absolute values and variability were observed (54.92 ± 39.36 data points, two-sample t-test: ****p < 10^−5^). (E) Result from a test run of the algorithm. The iteration counts were plotted as a function of the level series. Detection of time-locked responses (L ≤ 2 data points) required more iterations of averaging at suprathreshold (black dots, 1.40 ± 1.06, mean ± s.d.) than subthreshold stimulus levels (cycles, 7 ± 0, mean ± s.d.; two-sample t-test, ****p < 10^−5^). After two consecutive hits at the iteration upper limit (N = 7, dash line), the algorithm flags no response for the applied stimulus level (cycles) and triggered a stop command to avoid nonproductive attempts with weaker stimuli. The algorithm-determined threshold matches visual identification.

Next, to test whether the algorithm could find the hearing threshold in real time, we grouped 60 sweeps as a batch then increment the group averaging by iterations (multiply by 60 for end sweep number) until positive results, which were scored by triple cross-correlation analysis, were returned by the algorithm. As shown in **Figure 2E**, the normalized count was small at the suprathreshold levels, while increased rapidly at near-threshold levels and reached its upper limit at subthreshold levels. Therefore, the estimated threshold was defined above the stimulus level at which the iteration upper limit was reached (the highest subthreshold level). Further attempt was made to model the change in iteration counts at near-threshold levels. We acquired an ABR dataset with 1-dB step size (**Figure S1A**) and fitted with both exponential and sigmoidal functions (**Figure S1B**). The visually identified threshold was found approximately at the stimulus level which yield 1.0 and 0.9 on the best-fitted exponential or sigmoidal function. In addition, we found the algorithm outcome was not strongly influenced by either the allowed lag for response detection (**Figure S2A**) or the applied iteration upper limit (**Figure S2B**) in the cross-correlation analysis, suggesting that the threshold determination is robust and does not require fine parameter tuning.

### ABR Threshold Determination in Human

To test whether the automated approach applied to human ABR, we acquired ABR datasets from eight human participants. Because intermediate single sweeps are not available on the commercial device, alternatively we provided the algorithms with recorded average responses over different sweep numbers (see **METHOD DETAILS**). Example average responses (**Figure 3A**), as well as group averages (**Figure 3B**), are shown at tested stimulus levels (60 dB SPL to 0 dB with a step size of 5 dB). The obtained CC peak amplitudes and the signal lags are plotted as a function of the stimulus levels (**Figure 3C** and **3D**). Note that 500 sweeps were added per iteration; the upper limit of iteration count was set to seven (3500 sweeps) and a larger decision boundary of lag (≤ 6 data points or less than ± 0.3 ms) for positive responses was implemented due to broader waveforms in human ABR evoked by click-sound. As shown in **Figure 3E**, the iteration count increases fast to reach the upper limit near the visually identified threshold level.

**Figure 3.**
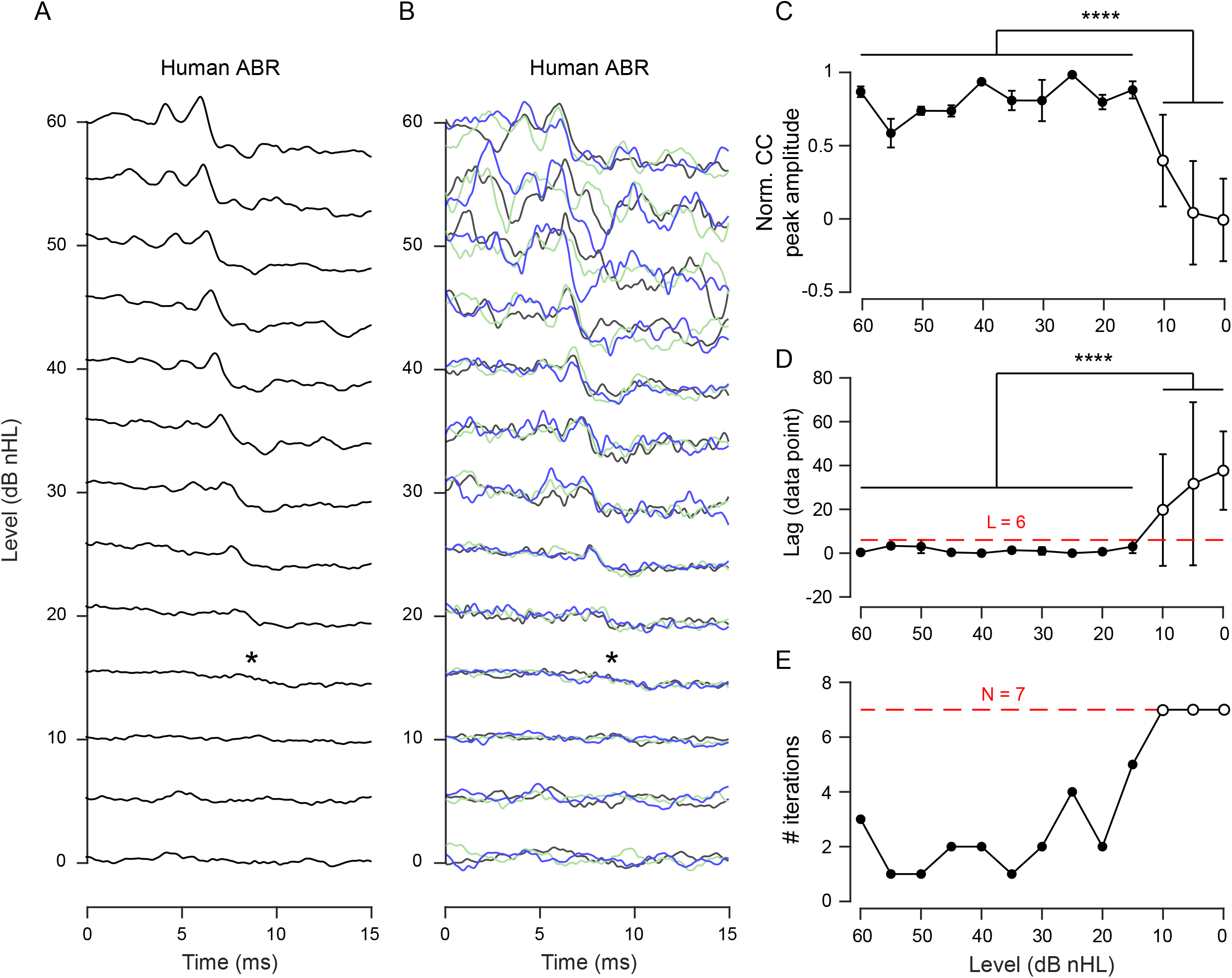
Threshold determination of human ABR by cross-correlation analysis. (A) Example level representations (averaging over 3500 sweeps) of a human participant with the visually identified threshold at 15 dB (asterisk). (B) Group averages that used in the algorithm for computing the cross-correlation. (C) Plot of the obtained CC peak amplitudes versus the level series. (D) Plot of signal lag at the CC peak versus the level series. At suprathreshold levels (dots, ≥ 15 dB), small mean value of the signal lags was obtained (1.30 ± 1.84 data points, mean ± s.d.), whereas the subthreshold level (< 15 dB, cycles) features a significantly large and variable lag value (23.00 ± 24.93 data points, two-sample t-test, ****p < 10^−5^). (E) Result from an algorithm test run. The iteration counts were plotted as a function of the level series. Detection of the true responses by the cross-correlation analysis (L ≤ 6 data points) within the iteration upper limit (N = 7, dash line) flags the suprathreshold levels (dots), which is consistent with those identified visually.

### Comparison between Expert and Algorithm Determined Thresholds

In order to evaluate the performance of our method, we recruited five human experts to assess the same ABR sets independently and compared their readouts of the thresholds to the algorithm outcomes (**Table 1**). Both approaches reached similar conclusions for both mouse and human ABR (**Figure 4A** and **4B**), validating reliability of the algorithm. Moreover, averaging over varied sweep numbers as used in the algorithm does not affect the threshold determination by either machine or human (**Figure S3A** and **S3B**). By further quantifying the minimum sweep number required by the algorithm (comparing results from the fixed number for all stimulus levels, 420 for mouse and 3500 for human ABR), we concluded that the algorithm avoided 66.72 ± 4.98% and 43.19 ± 12.48% of nonproductive sweeps, respectively (**Figure 4C** and **4D**).

**Figure 4.**
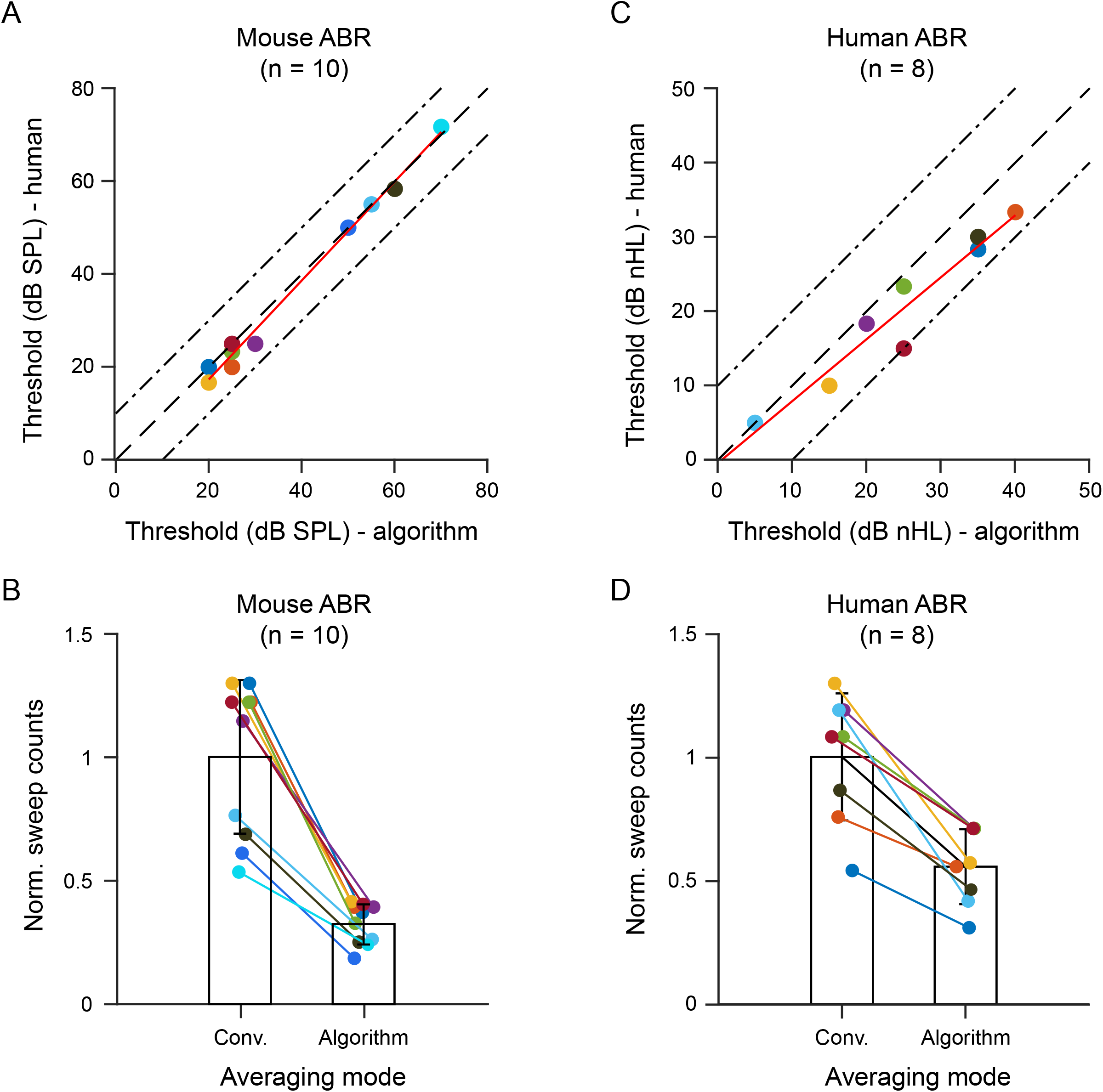
Comparisons between the thresholds determined by machine and human. (A) For mouse datasets, a close match was found between the algorithm-determined thresholds and those averaged from three out of five independent human readouts (maximum and mean discrepancies, 5 dB and 1.83 ± 2.45 dB, mean ± s.d.). Linear fit: adjust R^2^ = 0.99. (B) Comparison between the total sweeps used in the conventional level averaging (420 sweeps at each level, left bar) and in the algorithm (with varying numbers, right bar). The latter requires 66.72 ± 4.98% (mean ± s.d.) fewer sweeps. Note that the sweeps were counted from all suprathreshold and two successive sub-threshold levels. (C) As for human datasets, similar thresholds were reported by both the algorithm and human readout (maximum and mean discrepancies, 10 dB and 4.58 ± 3.88 dB, mean ± s.d.). Linear fit: adjust R^2^ =0.93. (D) The total number of sweeps used in the conventional level averaging (left bar) versus in the algorithm (right bar). The latter requires 43.19 ± 12.48% (mean ± s.d.) fewer sweeps.

## Discussion

Over decades several statistical approaches have been proposed to automatically detect ABR waveforms. Cross-correlation is one of the most favorite methods, as it can detect temporally stable waveforms with high sensitivity and robustness in a template-free fashion. This is crucial for recognizing ABRs, which oftentimes present large intersubject variability in terms of waveform shape and latency, from different levels of background noise. However, prior attempts using this approach relied on arbitrary decision boundary for response detection, for instance, minimum required CC (Berninger et al., 2014; Bershad and Rockmore, 1974; Suthakar and Liberman, 2019; Weber and Fletcher, 1980) or maximum allowed latency shift (Galbraith and Brown, 1990; Xu et al., 1995). Preselection of such criteria can be problematic in practice because even for the same test subject, comparable SNR across recording sessions is not guaranteed due to variabilities in electrode impedance and placement, as well as level averaging settings preferred by individual experimenter. Thus, it is unlikely that a universal response decision boundary can be applied on all ABR sets without introducing detection error, barring its usage in cases like cross-institution collaboration efforts where data pooling is needed.

Our approach, which is fundamentally different from the existing approaches, determines the hearing threshold based on the relative change in the sweep numbers required for positive ABR detection at different stimulus levels by the cross-correlation. The resulted signal lag of CC peak has proven to be a reliable criterion for detecting the time-locked responses with high sensitivity, but in principle, other quantifications like CC peak amplitude (**Figure 2C**) or single-point F-distribution (data not shown) can also be used in the algorithm. At near-threshold levels, a rapid increase in the end sweep number was observed (**Figure 2E** and **3E**). It is not surprising because in order to reach confidant SNR, small responses require increased averaging for baseline noise reduction, which means that the end sweep numbers quantitatively noted by the algorithm reflect the relative change in the SNR at different recording levels. Besides, this approach does not heavily rely on the selection of detection parameters. First, sub- and supra-threshold level representations feature large differences in the obtained signal lags (**Figure 2D**), which offers a wide range of decision boundary selection of lag without affecting detection accuracy (**Figure S2A**). Second, raising upper limit of the end sweep number only leads to a small shift from the estimated threshold (**Figure S2B**) due to the exponential increase of averaging at near-threshold levels (**Figure S1B**),

So far, the precision of threshold determination is constrained by the step size of level sampling. Modeling the obtained end sweep number change (**Figure S1**) and interpolating the level representation may produce higher precision. Further development of this approach is to combine with level sampling strategy including progressively reduced step size (Cebulla and Sturzebecher, 2015) and increased batch size of sweeps per iteration at near-threshold levels so that the model fitting can be improved by more effective data points in the transition.

In mouse ABR, the proposed method was proven reliable in threshold determination with a mean difference of 1.84 dB (n = 10) and maximum discrepancy of ± 5 dB between the algorithm outcomes and human readouts (**Figure 4A**). Although the detection accuracy is comparable to that of the most up-to-date approach (Suthakar and Liberman, 2019), our algorithm has the advantage of not requiring a calibrated preset criterion and has more robust outcome. Validation with human ABR resulted in only slightly worse detection accuracy (mean and maximum difference, 4.58 and ± 10 dB; **Figure 4C**) than that of mouse ABR. Moreover, the ABR sets with average responses over varying sweep numbers as in the algorithm do not seem to introduce additional difficulty for visual threshold determination (**Figure S3A** and **S3B**). This is not surprising because excessive level averaging at suprathreshold levels is not expected to improve the threshold determination. This feature is extremely attractive for two reasons. First, it provides minimal quality control for unambiguous waveform recognition for both human and machine. Such standardized data collection will benefit artificial-intelligence based approaches by improving the quality of training data (McKearney and MacKinnon, 2019). Second, when to terminate averaging without compromising the quality of the recording is an important decision making during ABR recording (Don and Elberling, 1996; Madsen et al., 2018), this method has the potential to instruct the efficient test by avoiding nonproductive recordings at supra-threshold levels.

### Limitations of the study

This is a proof-of-principle study for detecting time-locked responses in cumulatively averaged level representations using cross-correlation analysis. As level averaging is terminated earlier for supra-threshold levels, the algorithm should be able to shorten the test duration. However, quantification of the exact saved time in a real setting requires implementation of the algorithm on an optimized hardware, which is currently under engineering.

## Data Availability

All data referred to in the manuscript are available.

## Acknowledgments

We thank Dr. Hao Wu for the support of this study. We thank Drs. Guangming Chen and Lin Liu for contributing terc^-/-^ mice. We thank Eric Song for comments on the manuscript. This study was supported by Shanghai Huangpu District Industry Support Fund (XK2019011 to Y.H.), the National Science Foundation of China (81770995 to L.S., 81700903 to B.L. and 81800901 to Y.H.) and the Shanghai Key Laboratory of Translational Medicine on Ear and Nose diseases (14DZ2260300).

## Author contributions

Y.H. designed the study; Y.H. and L.S. supervised the study; B.L., K.H., X.W. and Z.W. contributed the human ABR datasets; Y.L. and Y.Q. performed the mouse ABR recording; H.W. and Y.H. wrote the algorithm and analyzed the data with the help of Y.L., Y.Q., H.S., J.Z. and L.S.; Y.H. wrote the manuscript with the help of H.W. and L.S.

## STAR METHODS

### RESOURCE AVAILABILITY

#### Lead Contact

Further information and requests for resources and reagents should be directed to and will be fulfilled by the Lead Contact, Y.H.

#### Materials Availability Statement

This study did not generate new unique reagents.

#### Data and Code Availability Statement

The codes were written in MATLAB scripts and included in the supplementary materials. All single sweep ABR datasets supporting the current study will be made available upon publication: https://data.mendeley.com/datasets/4yb9772dff/draft?a=d2d509bc-2a09-426b-a1c8-25ef9c817455

## EXPERIMENTAL MODEL AND SUBJECT DETAILS

C57BL/6 and CBA mice were purchased from Sino-British SIPPR/BK Lab Animal Ltd. (Shanghai, China). The telomerase-knock-out mice (terc^-/-^) were gifts from Prof. Lin Liu (Nankai University, China) and bred in house. Human participants were recruited by Hearing and Speech Center, Shanghai Ninth People’s Hospital and consent forms were signed before the experiment. This study was conducted at the Ear Institute and the Hospital Hearing and Speech Center. All procedures were reviewed and approved by the Institutional Authority for Laboratory Animal Care (HKDL2018503) and the Hospital Ethics Committee for Medical Research (SH9H-2019-T79-1).

## METHOD DETAILS

### ABR Recording

Mouse ABRs were recorded via a TDT RZ6/BioSigRZ system (Tuck-Davis Tech. Inc., US) in a sound-proof chamber as previously described (Lin et al., 2019). In brief, 7-week-old animals were anesthetized through intraperitoneal injection of Chloral hydrate (500 mg/kg). During the recording, animal body temperature was maintained at 37°C using a regulated heating pad (Harvard Apparatus, US) with a rectal thermal probe placed under the body. Evoked potentials were registered via subdermal needle electrodes (Rochester Electro-Med. Inc., US) placed at the animal’s vertex (active electrode), left infra-auricular mastoid (reference electrode) and right shoulder region (ground electrode). 3-ms tone pips at 16 kHz were delivered via an MF1 speaker (Tuck-Davis Tech. Inc., US) positioned in the front of the animal 10 cm away from the vertex. Acoustic stimuli were presented 20 stimuli per second and the evoked potentials were collected at 24 kHz sampling rate. Artifact rejection level was set to < 35% (mean rejection voltage 20.5 μV). Sound level series started from 90 to 0 dB sound pressure level (SPL) with 5-dB step size. For one animal, the stimulus level series were repeated from +10 to –10 dB SPL around the estimated threshold with 1-dB step (**Figure S1**).

Human ABRs were recorded by a commercial ABR device (Intelligent Hearing Systems, US) with Smart EP software from eight volunteers aged 23-73 years without the knowledge of their medical conditions. Sound stimulation (100 μs duration, rectangular envelopes) was generated and presented monaurally through ER3 insert earphones with foam tips. Stimuli were presented at a rate of 37.1/sec with alternating polarity. Electrode impedance was < 5 kΩ and inter-electrode impedance was within ± 1 kΩ. The artifact rejection level was < 31% (rejection voltage 31 μV) to exclude contaminations from EEG and myogenic potentials. The evoked potentials were collected with 20 kHz sampling rate and ×100,000 amplification. The bandpass filter was set at 100-3000 Hz. Average responses over 500, 1000, and 2000 sweeps were acquired and repeated three times for the level series starting from 60 to 0 dB SPL with 5-dB step size.

### Cross-correlation Analysis of Mouse ABR

Sweeps were loaded in three memory buffers. Cross-correlation (MATLAB Central File Exchange Function *xcorr*, MathWorks, US) was computed from two out of three group averages. This resulted in correlation coefficients as a function of signal lag between two group averages). As the ABRs were time-locked to the stimulus onset, they could be detected based on a neglectable signal lag where the maximal coefficient was found (Xu et al., 1995). In this study, maximum allowed lag (*L*) for a true ABR signal was set to ± 2 data point from time zero (equivalent to ± 0.082 ms or 1% of the analyzed temporal window). Three such tests were implemented in parallel in order to minimize false positives caused by coincidently overlapped background activities through rejecting inconsistent lag values. Moreover, the obtained correlation coefficient peak amplitude was included as an independent variable (**Figure 2C**).

The test started with the loudest stimulus level (90 dB SPL) and was reduced by a step size of 5 dB. At each given stimulus level, the sweep number used to produce the group average was increased by iterations and the end sweep number was noted upon detected response determined by the cross-correlation analysis. For each iteration, sweeps were added in batches (60 sweeps) and an upper limit was set (420 sweeps) to avoid nonproductive attempts at subthreshold levels. Upon detected responses, the iteration was then switched to lowered stimulus level, until the upper limit was reached for two consecutive nonproductive trails. The estimated threshold was the lowest applied stimulus level at which the upper iteration limit was not yet reached.

The transition of the end sweep numbers between supra- and subthreshold levels was modeled (**Figure S1**). Both sigmoidal (1) and exponential functions (2) were employed to fit the relationship between the normalized iteration count *C’* (equivalent to the sweep number) and the stimulus level *S* using a nonlinear least square method in MATLAB (Curve Fitting Toolbox, MathWorks, US). In the functions, α_1_ = 0.6 and α_2_ = 0.2 were fixed for calibrated lag criterion (*L* = 2), whereas β_1_ and β_2_ were obtained by fitting.

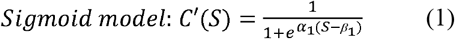

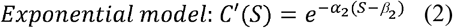

### Cross-correlation Analysis of Human ABR

For human ABR, group average responses were recorded sequentially and used directly as inputs of the algorithm. Group averages over 500, 1000 and 2000 sweeps were recorded, whereas averages over 1500, 2500, 3000 and 3500 sweeps could be obtained by linear combination (3) where *E*{*m*}, *E*{*n*} and *E*{*m* + *n*} denote the time averages over *m, n* and *m* + n sweeps, respectively.

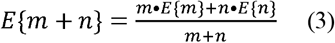

The maximum allowed lag for a true response was set to ≤ 6 data points from time zero (equivalent to ± 0.3 ms or 2% of the analyzed temporal window). The iteration upper limit was seven, corresponding to 3500 sweeps. The estimated threshold was the lowest stimulus level with a detectable response.

### Threshold Determination by Human Readout

To estimate the ground-truth thresholds of the recorded mouse and human ABRs, average responses of all level series were provided to five clinicians to report the visually identified thresholds independently. The test subject identities were blinded to the judges. Either the fixed sweep number (the conventional averaging) or the varying numbers used in the algorithm (the algorithm averaging) was applied to compute the level averaging. The thresholds were determined by three out of five execution judges (with the highest and the lowest value excluded). The readouts were used to evaluate the accuracy of the algorithm outcomes (**Table 1**).

## Table titles and legends

**Table 1.** Comparison between algorithm outcome and human readout

---

**Figure S1.**
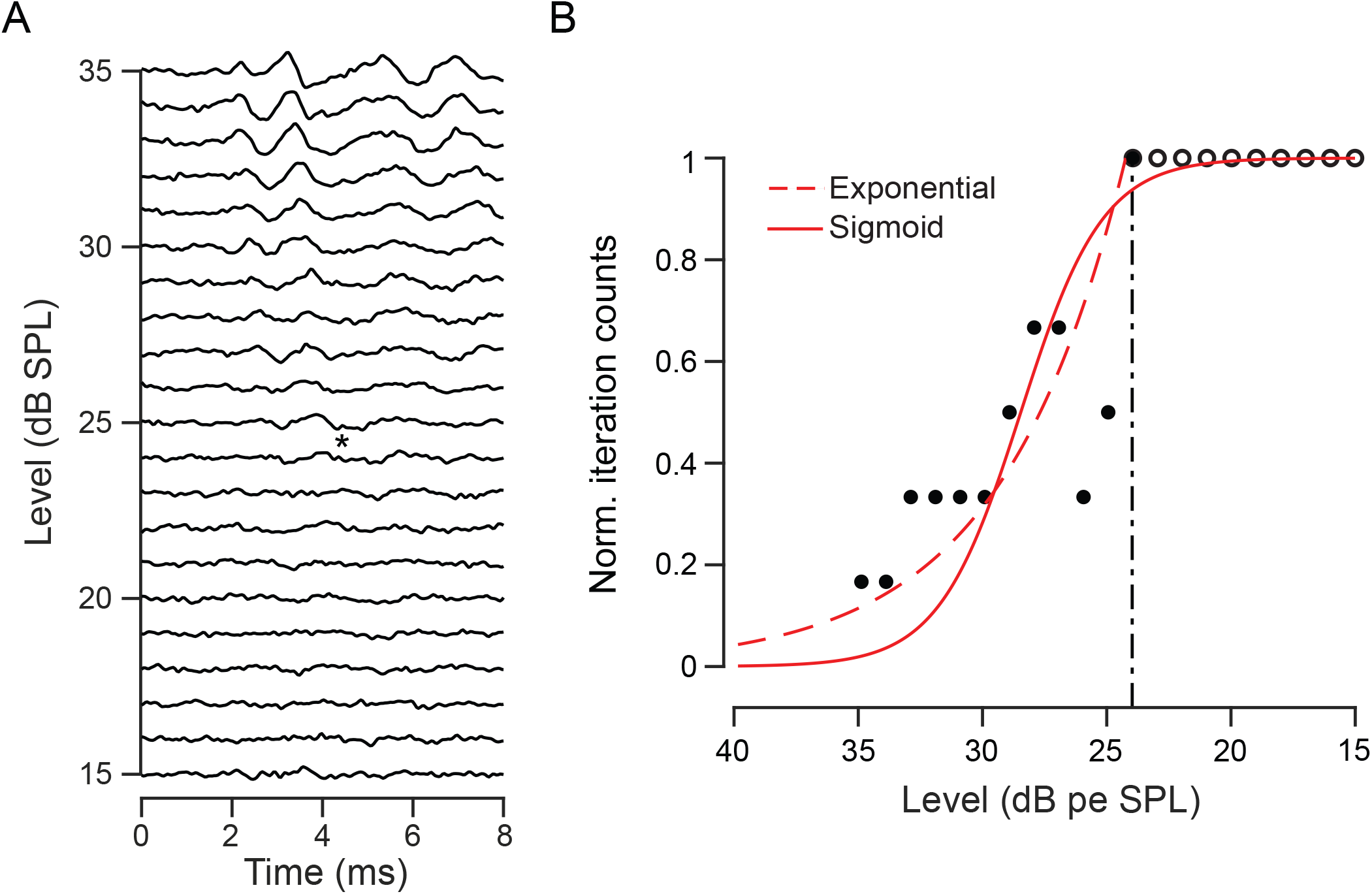
Modeling of the change in the algorithm iteration counts. (A) Average level representations acquired from a mouse by stimulations with a step size of 1 dB. The visually identified threshold was 24 dB (asterisk). (B) The normalized iteration counts versus level series is fit by both sigmoid (solid line) and exponential functions (dash line). The visually identified threshold is approximately at the level corresponding to 1.0 and 0.9 of the best-fit exponential and sigmoid growth, respectively.

**Figure S2.**
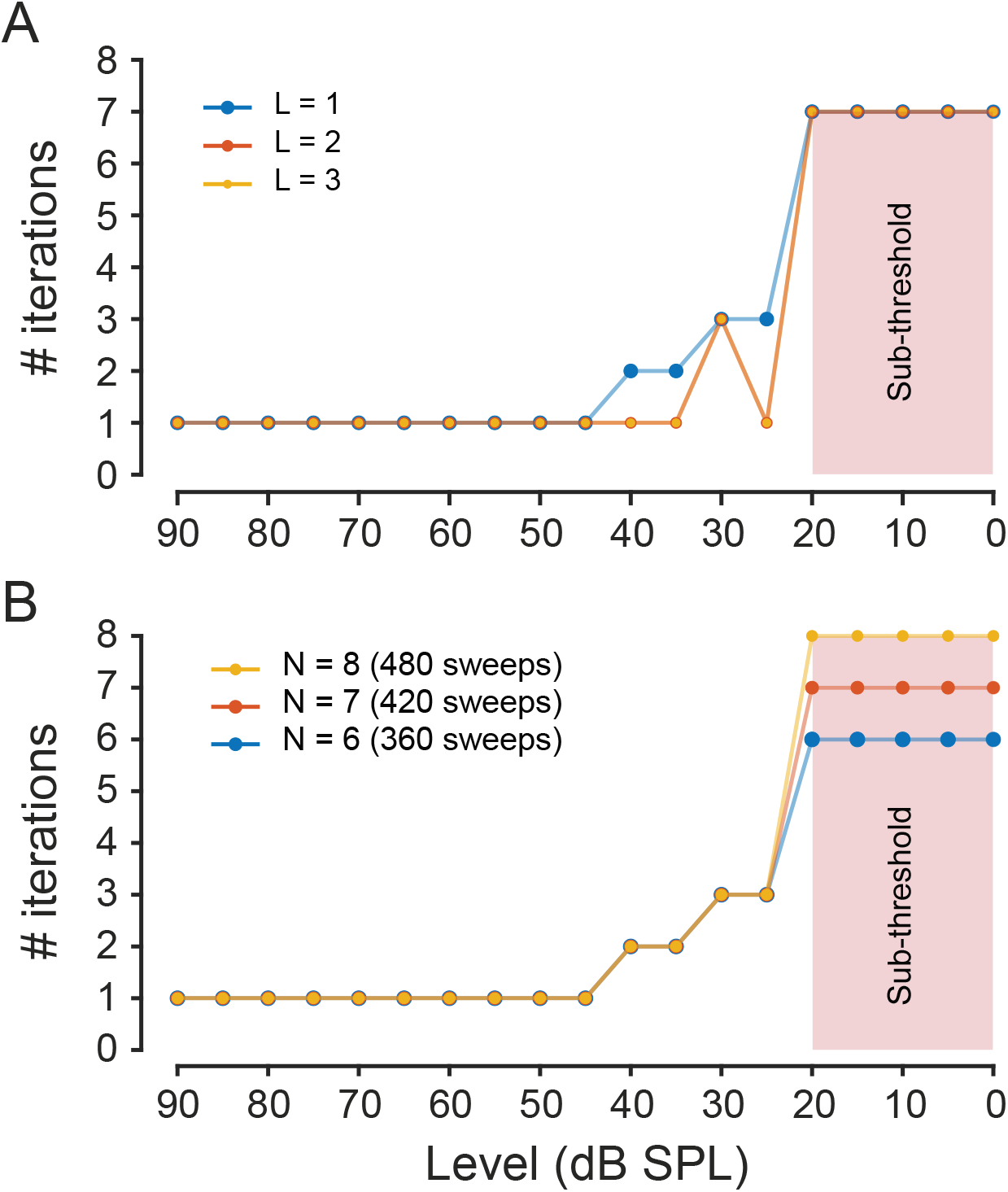
Selection of detection parameters has limited influence on the threshold determination. The algorithm outocmes of allowing different signal lags for ABR detection on the same mouse dataset. Despite of changes in the iteration counts, the resulted threshold did not drift. (B) The algorithm outcomes of using different iteration count upper limit. The sub-threshold levels flagged by reaching the upper limit were not altered.

**Figure S3.**
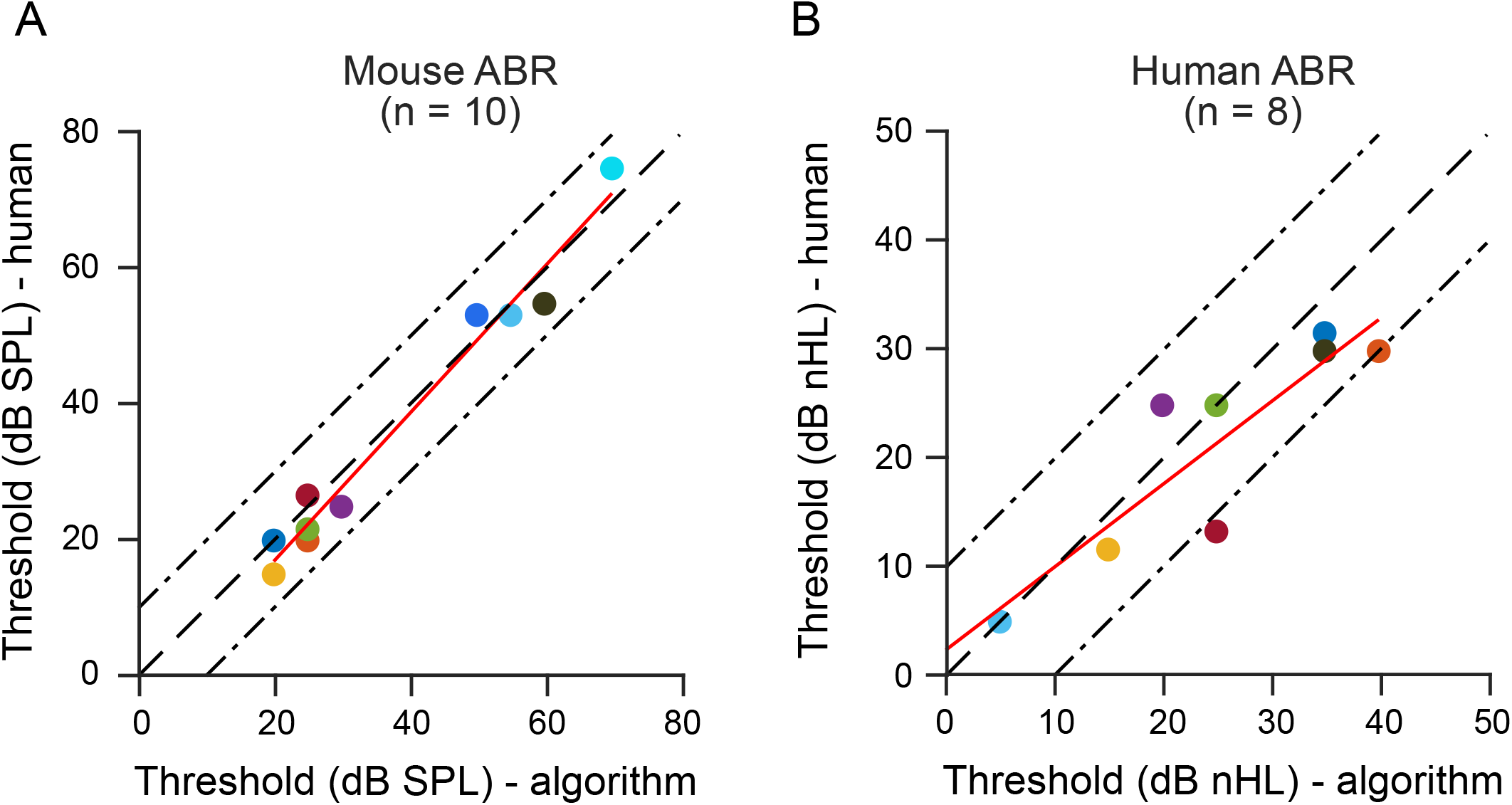
Varying level averging used in the algorithm does not affect the threshold determination by visual identification. (A) Level averages of mouse ABR datasets based on the algorithm iteration counts were provided to human experts to readout the threshold. The results were consistent to those determined by the algorithm (maximum and mean discrepancy, 5.00 dB and 3.50 ± 2.33 dB, mean ± s.d.). Linear fit: adjust R^2^ = 0.97. (B) Similar results were obtained from both human readouts and the algorithm on the level averages of human ABR dataset over varying sweep number (maximum and mean discrepancy, 15 dB and 5.21 ± 5.00 dB, mean ± s.d.). Linear fit: adjust R^2^ = 0.78.

